# Coccidioidomycosis Emergence in South America: Exploring Northeastern Brazil’s Epidemiological, Clinical, and Genomic Landscape

**DOI:** 10.1101/2023.08.14.23294078

**Authors:** Kelsen Dantas Eulálio, Daniel R. Kollath, Liline Maria Soares Martins, Antonio de Deus Filho, Maria do Amparo Salmito Cavalcanti, Lucas Machado Moreira, Bernardo Guerra Tenório, Lucas Gomes de Brito Alves, Danielle Yamauchi, Gil Bernard, George R Thompson, Mathieu Nacher, Jason E. Stajich, Eduardo Bagagli, Maria Sueli Soares Felipe, Bridget M. Barker, Luciana Trilles, Marcus de Melo Teixeira

## Abstract

Coccidioidomycosis is an invasive mycosis included in WHO’s priority list. It is endemic and notifiable in the United States but neglected in Central and South America. We used a multi-institutional approach to assess whether disease characteristics, genetic variation in the pathogen or environmental factors affects the epidemiology of coccidioidomycosis and disease outcomes throughout the American continent. We identified 292 patients with coccidioidomycosis between 1978 and 2021 in the Piauí and Maranhão states of Brazil; the largest cases series reported outside the US/Mexico epidemic range. The male-to-female ratio was 57.4:1 and the main risk factor was armadillo hunting (91.1%) 4 to 30 days before symptom onset. Forty-two outbreaks involving two to six patients were observed. Most patients (92.8%) presented typical acute pulmonary disease, followed by disseminated (3.4%), chronic pulmonary (2.4%) and regressive pulmonary (1.4%). The most frequent clinical symptoms were cough (93%), fever (90%) and chest pain (77%). Mortality was observed in 8% of the patients. In 2004, and between 2015 and 2017, we observed a spike in coccidioidomycosis in Brazil, particularly in the state of Piauí. Unlike other main hotspots, the soil is acidic in this region and precipitation (p=0.015) and precipitation one-year prior (p=0.001) were predictors of higher coccidioidomycosis rates. The Brazilian strains are genotypically divergent from other described *C. posadasii* within the Texas/Mexico/South America clade. Coccidioidomycosis in Northeastern Brazil has a specific infection profile and armadillo hunters are at higher risk. Low pluviosity and extensive drought appear key to increasing the number of cases in Brazil. A unique *C. posadasii* lineage exists in Brazil; therefore, environmental, virulence, and/or pathogenesis traits may differ from other *Coccidioides* genotypes.

## Background

Coccidioidomycosis (CM) is an invasive mycosis included in the WHO list of priority fungal pathogens. It is endemic and notifiable in the USA but is neglected in Latin America^1^. The disease caused by *Coccidioides immitis* is limited to California, Washington, and Northwestern Mexico, whereas *C. posadasii* infections are broadly distributed in western USA and Latin America^2^. *Coccidioides* spp. are saprotrophic dimorphic fungi phylogenetically placed in the order Onygenales (Pezizomycotina, Ascomycota)^3^. This fungus produces infectious arthroconidia in the environment that are aerosolized upon soil disturbance. Mammals, including humans, inhale arthroconidia, which are phagocytosed by alveolar macrophages triggering a morpho-physiological change to pathogenic spherules^4^. In endemic areas, the distribution of the fungus in the soil is focal and heterogeneous, and animal dens and archaeological sites represent the most common positive sites^5,6^. The number of infections in California and Arizona ranks disease burden as one of the highest reported infectious diseases, with high morbidity and mortality. It is estimated that over 500,000 people are naturally infected per year in the US and increases in cases have been observed in association with severe droughts^7,8^. Approximately 60% of *Coccidioides* infections are asymptomatic and symptomatic cases are primarily mild and may be confused with other causes of community-acquired pneumonia. However, some patients experience chronic illness with symptoms lasting several months to years. Fewer than 5% of symptomatic cases develop into severe and disseminated forms or life-threatening disease ^9^.

Coccidioidomycosis is endemic to arid and semi-arid hotspots in Central and South America but with a lower incidence than North America. Interestingly, 100 years after the first report of the disease in Argentina, fewer than 1,000 total cases have been reported^1^. Coccidioidomycosis is not a notifiable disease in Latin America, so true incidence has not been determined. We hypothesize that altered virulence traits, lack of diagnosis and reporting, and bioclimatic and demographic variations in Latin America could explain the lower incidence compared to North America^1,2,7^. Northeastern Brazil has been recognized as an endemic area since 1998, but initial case reports date to the 1970s ^10-12^. In Brazil, *Coccidioides posadasii* has been isolated from the environment, humans, dogs, armadillos, and bats^12,13^; however, the true coccidioidomycosis burden in Brazil is unknown. Only a few cases, with scarce epidemiological data, have been reported in Piauí, Ceará, Maranhão, Pernambuco, and Bahia states^12^, all northeastern states with vast semi-arid areas.

Given the lack of knowledge, we designed a comprehensive assessment of coccidioidomycosis in Brazil. Our unbiased and multi-institutional approach to assess genetic variation in the pathogen and environmental components that might affect the epidemiology of coccidioidomycosis. Therefore, we 1) described the largest case series study of coccidioidomycosis to date in South America from Piauí and Maranhão states of Northeastern Brazil; 2) used geographical coordinates related to clinical, veterinary cases, and environmental detection coupled with bioclimatic and demographic factors to predict the true geographic range of the disease in Brazil; and 3) inferred the genotypes of *C. posadasii* in Northeastern Brazil using whole-genome analysis. This study will increase awareness of this severe mycosis in South America; and exploring coccidioidomycosis outside California and Arizona deepens our understanding of the species complex disease dynamics.

## Methods

### 1 Clinical data

We conducted a retrospective study of microbiologically proven coccidioidomycosis cases between 1978 to 2021 at the Institute of Tropical Diseases Nathan Portela and Pulmonology Clinic of Hospital Getúlio Vargas. Patients showing clinical respiratory signs were evaluated and inclusion criteria were based on the isolation or observation of fungal cells compatible with *Coccidioides* (Appendix 1). Lung damage was assessed using chest X-ray and/or computed tomography (CT). Residual lesions such as nodules or cavitation and sequelae such as calcifications, fibrosis or bronchiectasis were investigated. Demographic data and risk activities were recorded. In cases of exposure to dust from armadillo habitats, the participation and eventual illnesses of the dogs were also investigated and managed accordingly (Appendix 1). The cases were classified as microepidemics when two or more human cases were diagnosed with coccidioidomycosis signs and symptoms that appeared simultaneously after a shared exposure event. Clinical signs, symptoms, and antifungal treatment of 100 patients with laboratory-proven coccidioidomycosis were retrieved through review of medical records and prospectively through subsequent clinical reviews.

### 2 Georeferencing, species niche modelling and environmental factors associated with coccidioidomycosis in Brazil

We collected patient addresses from our cohort and used previously published environmental data, including *Coccidioides* spp. isolation from armadillos and positive soil samples, to georeference the entries using QGIS 3.10.9 software (http://www.qgis.org/) with SIRGAS 2000 Datum and a cartographic base from Brazilian Institute of Geography and Statistics (IBGE 2020). The species modeling distribution was assessed using the Maximum Entropy (Mexent) model, and negative-binomial regression tested which climate variables were related to the increase of cases in Brazil (Appendix 1)

### 3 Fungal isolation, genome sequencing and whole-genome genotyping

Fungal growth was achieved by plating clinical specimens (Appendix 1) into the fungibiotic media Mycosel. DNA was extracted from 500mg of cells, assessed for integrity via agarose gel electrophoresis, 1μg of input DNA subjected to libraries preparation using the NEBNext® Ultra™ II DNA Library Prep Kit (New England Biolabs) and quantified via qPCR, Bionalyzer (Agilent) and Qubit (Invitrogen). DNA libraries were sequenced using the NovaSeq 6000 instrument (kit v1.5 - 300 cycles - 2×150bp) in a high-throughput mode. SNPs were retrieved after a series of *in silico* genome alignments and filtering steps, phylogenomic analysis were performed under Maximum Likelihood criteria and Principal Coordinate Analysis (PCA) was performed to verify the population distribution within *C. posadasii*. Nucleotide diversity was calculated within each *C. posadasii* population (Appendix 1).

### 4 Ethical and regulatory aspects

This study was approved by the Research Ethics Committee of the Institute of Tropical Diseases Nathan Portela, subsequently ratified by the Ethics Committee of the Federal University of Piauí (No.36/08 - CAAE 0036.0.045.000-8).

## Results

### 1 Identification of risk factors for coccidioidomycosis in northeastern Brazil

We identified 292 patients with proven coccidioidomycosis between 1978 and 2021. Most (n = 270, 92.5%) were from the state of Piauí, with 77 (34.4%) of the 224 municipalities of the state contributing cases. Reported cases span both caatinga and cerrado biomes, particularly the semi-arid cerrado-to-caatinga transition. Table 1 contains patient clinical and demographic characteristics. Males were predominantly affected (male to female ratio: 57.4:1, p < 0.0001). As a reflection of the predominant African-American background of Brazilian Northeastern region, most patients were also of this ancestry (78.4%) compared with Caucasians (20.9%, p=5.9E-23). Of note, there were only two indigenous patients (0.7%), both from the Guajajara ethnic group in the state of Maranhão.

**Table 1.**
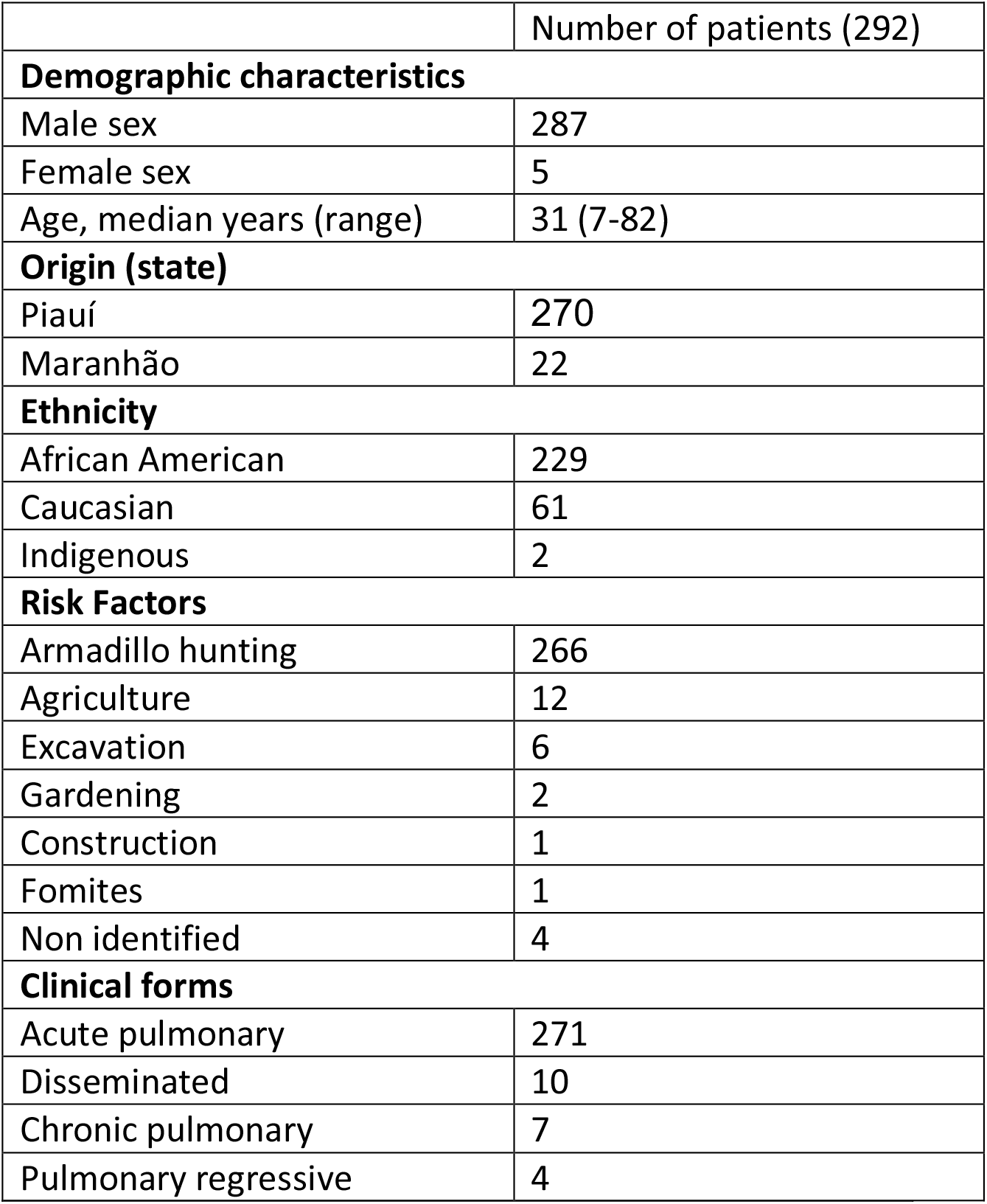
Demographic characteristics, risk factors and clinical forms of coccidioidomycosis in Brazil.

Armadillo hunting (n = 266, 91.1%) occurring 4 to 30 days before the onset of coccidioidomycosis-related symptoms was the primary risk factor identified and is unique to this area of Brazil (Table1, Fig. 1). Farming was the leading occupation (167/57.2%) followed by students (57/19.5%), trade workers (12/4.1%), and a range of other activities. The most affected age group was 20-39, corresponding to ∼50% of the patients; overall 72.6% were under 40 years-old.

**Figure 1.**
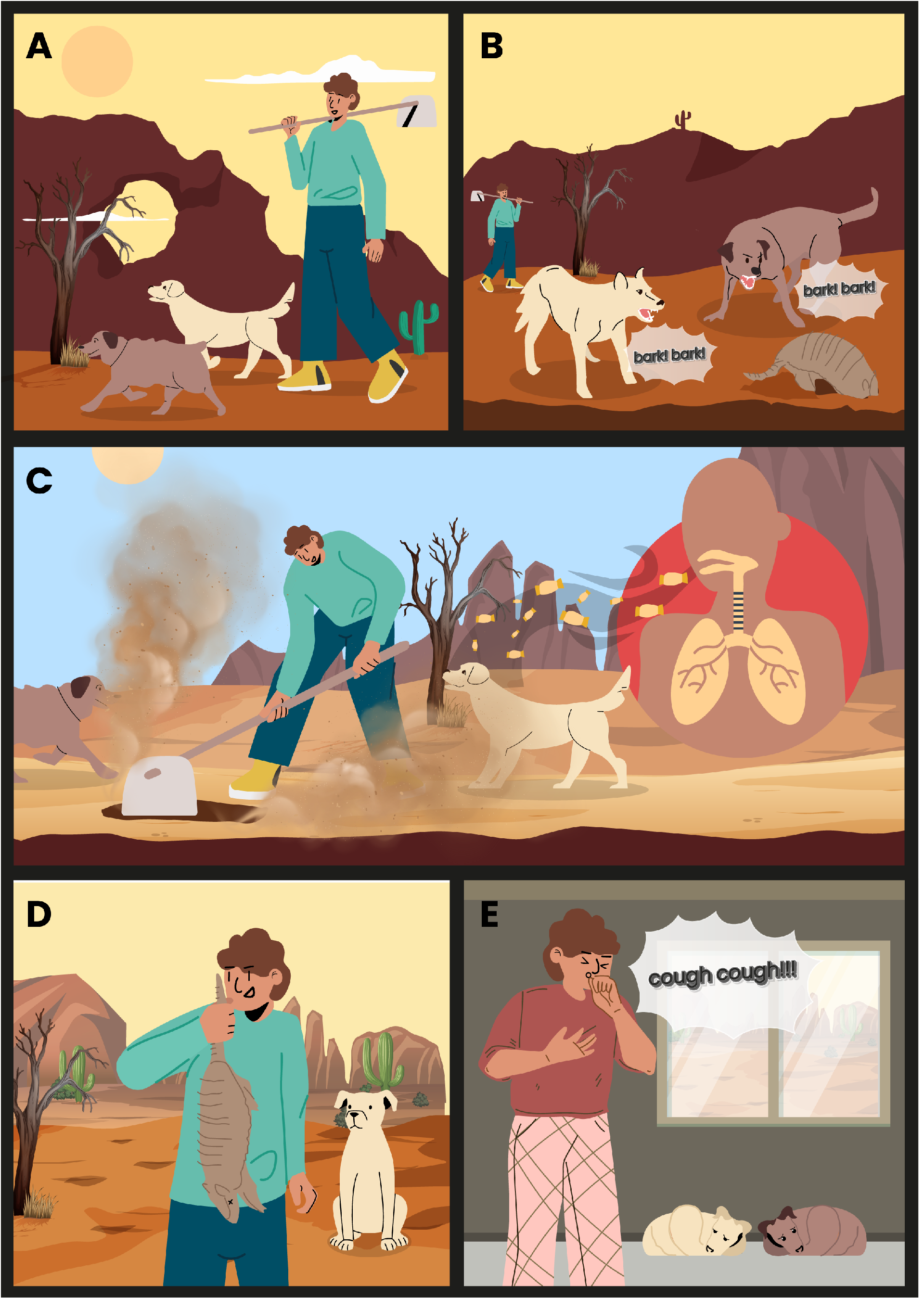
Coccidioidomycosis acquired during armadillo hunting practices in Northeastern Brazil. A) Armadillo hunting is a centenary practice in Brazil that use speed chasing and ambush by hunters and dogs, and is the most common risk factor for coccidioidomycosis in the Brazilian semiarid region. B) Hunting dogs smell the armadillos hundreds of meters away and chase the armadillos, which digs into its burrow to escape. The hunting dogs start to bark, alerting hunters. C) Once located, the hunter excavates the armadillo from its burrow with help of a hoe and shovel. Soil particles become aerosolized, and a large volume of dust is formed potentially containing infectious arthroconidia that can be inhaled by humans and dogs. D) After a lengthy excavation, the armadillo is finally extracted from its burrow and take to the hunter’s home to be consumed. E) Around 30 days after exposure, both humans and dogs develop the initial symptoms of acute coccidioidomycosis, such as cough, fever and chest pain.

Coccidioidomycosis occurred as single cases in 63.4% (n=185) of the instances and as microepidemics (two to six patients, average: 2.5) in 107 instances (39.6%). Microepidemics were related to armadillo hunting of both *Dasypus* sp. and *Euphractus* sp. in 41 of the 42 recorded micro-epidemics; in one case it was related to stone extraction for civil construction. Finally, of 65 dogs that participated in hunting, 23 developed coccidioidomycosis (35.4%), of whom eight died of the disease.

### 2 Acute pulmonary disease is the main form of coccidioidomycosis in Brazil

Patients’ clinical classification was primarily acute pulmonary disease (Table 1; n=271, 92.8%). Ten cases (3.4%) with an initial acute pulmonary disease evolved to disseminated disease. Seven cases (2.4%) presented with chronic pulmonary disease and 4 cases (1.4%) had residual pulmonary form. More detailed clinical and radiology data were collected from a subset of 100 patients (Table 2). The most frequent symptoms were cough (93%), fever (90%) and chest pain (77%). Cough was predominantly dry (65.6%), or initially dry and increasingly productive in 28 cases (30.1%). Typical CAP-associated productive cough was rare (4.3%). Dyspnea was reported by 51% of the patients and 12% had severe pneumonia with acute respiratory failure. Hemoptysis occurred in four cases (4%).

**Table 2.**
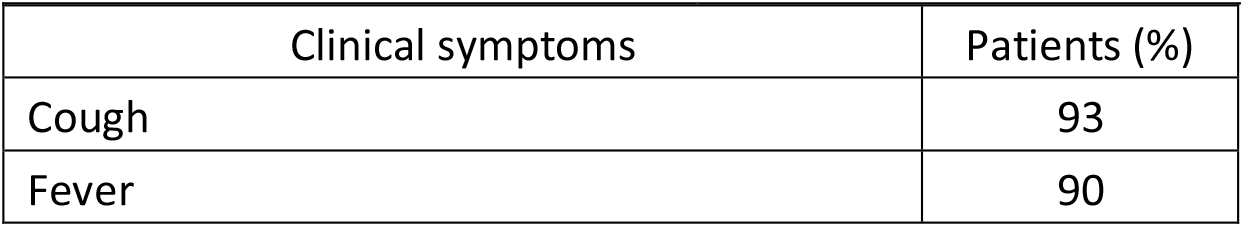
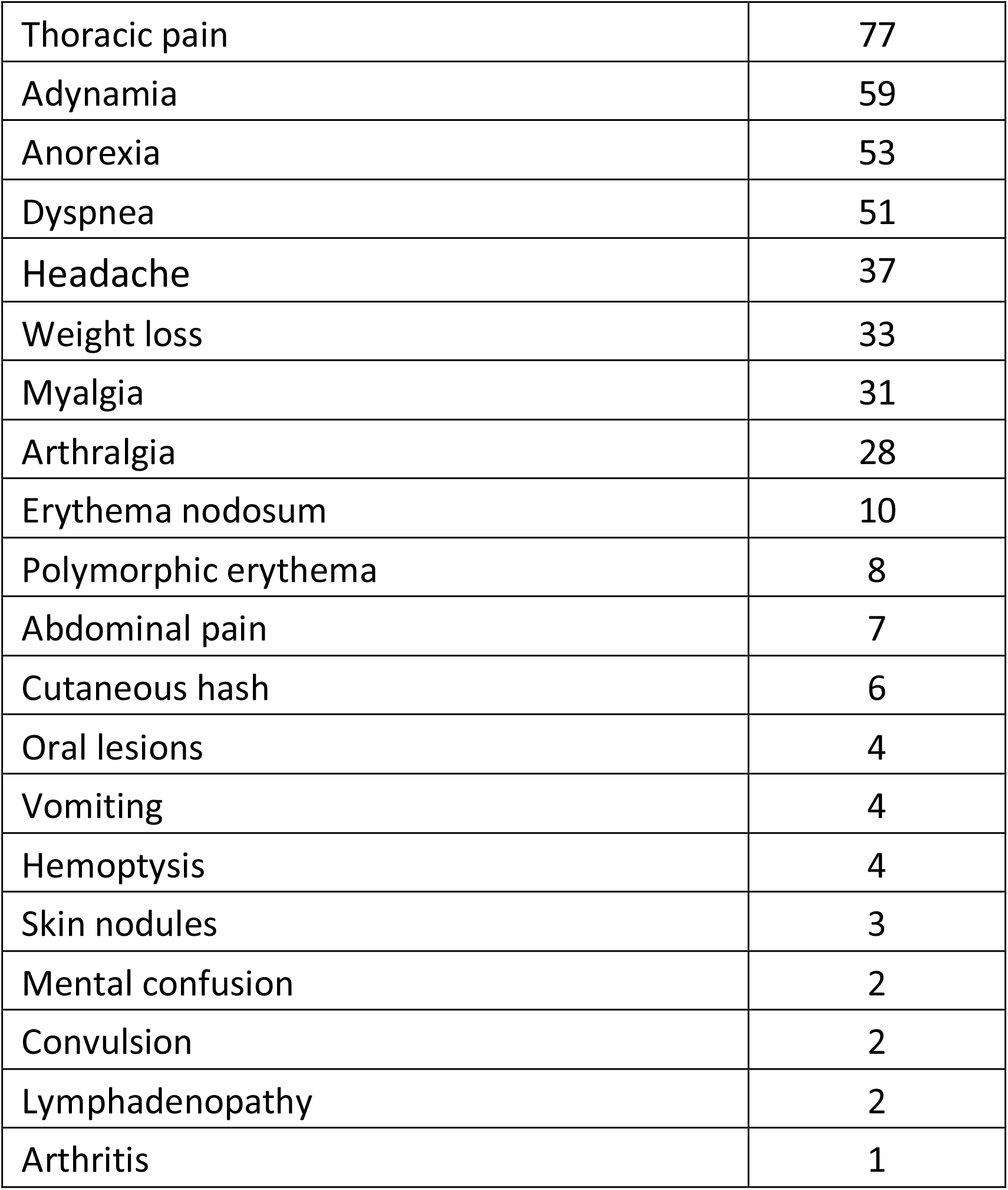
Clinical symptoms of patients diagnosed with coccidioidomycosis in Brazil.

Chest X-rays showed a bilateral multiple nodular pattern in 86.3% of patients. In 52.6%, this was the only radiological observation, 25.3% were associated with parenchymal consolidation, and 7.4% of patients had interstitial infiltration. Isolated parenchymal consolidation and interstitial infiltration were present in 10.5% and 3.2% of cases, respectively. The nodules were bilaterally distributed, predominating in lung base fields, measuring up to 3 cm in diameter, with ill-defined contours; excavation of the nodules was detected in 10.5% of the cases.

Dermatological manifestations of hypersensitivity were observed. Ten patients presented *erythema nodosum*, 8 *erythema multiforme* and 6 other types of exanthema; 4 patients had mixed manifestations (Table 2). Mucosal lesions were present in four patients with *erythema multiforme*. Among the 7 patients with disseminated disease, cutaneous/subcutaneous involvement was observed in 3, lymph node involvement in 2 and CNS involvement in 2 cases, one of the later with concomitant osteo-articular involvement. Azole antifungals were the most used medications alone (55%) or following deoxycholate amphotericin B therapy (24%): itraconazole was preferentially used (71%), followed by ketoconazole (6%), and fluconazole (2%). Deoxycholate amphotericin B alone was the therapeutic alternative used in 12% with severe manifestations. Treatment was effective and 92% of the patients resolved infection. However, eight patients evolved with spontaneous regression. Another eight patients (age range 15-63) died due to severe pulmonary involvement with respiratory insufficiency (n=6), involvement of CNS (n=1), or both (n=1).

Reassessment for post-treatment sequelae was carried out one to seven years (4.8±2.1) post anti-fungal therapy withdrawal in 25 randomly recruited patients, 24 with prior acute pulmonary disease and one with disseminated disease (cervical lymph node enlargements). Their age range was 13-51 years old. They were all asymptomatic and with no abnormalities on physical examination. However, thoracic imaging revealed residual pulmonary abnormalities in 92% of the evaluated patients. The main findings were multiple nodules (88%), calcified nodules (52%), and fibrosis (16%).

### 3 Severe droughts are key factors for increasing of coccidioidomycosis cases in northeastern Brazil

All cases of coccidioidomycosis were georeferenced, including the microepidemics, in Northeast Brazil. Beyond the Caatinga biome, which was extensively associated with the disease in Brazil, we have found cases in the Brazilian Savana (Fig. 2).

**Figure 2.**
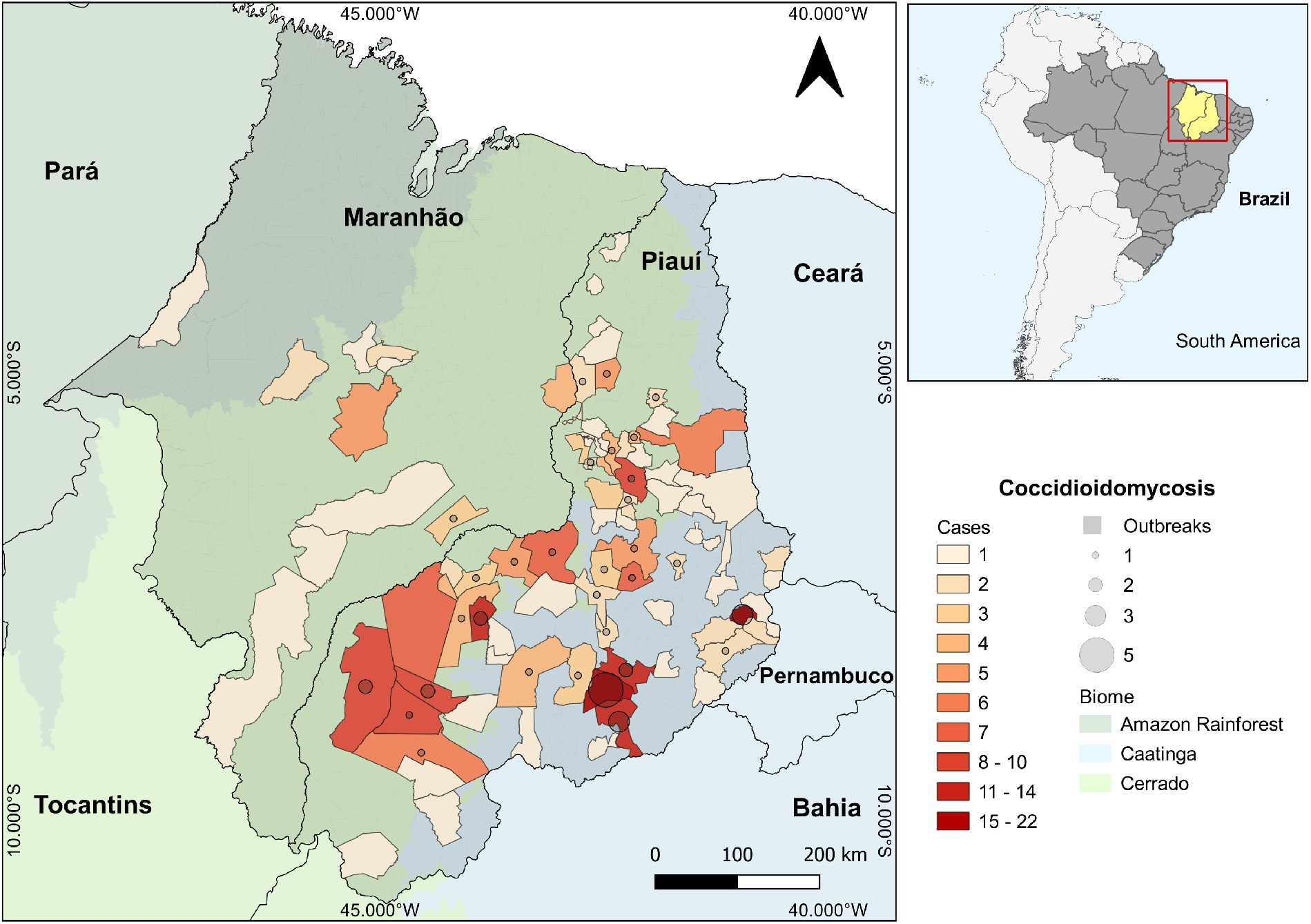
Distribution of coccidioidomycosis cases in northeastern Brazil. The 292 coccidioidomycosis cases in Brazil were georeferenced and assigned to each municipality in Piauí and Maranhão states; scale transitions from pale, the lowest number of cases, to dark red, the highest number of cases. Micro-epidemics are represented by circles within each municipality; the higher the number of episodes, the greater the size of the circles. The main biomes are also represented by map layers delimited by different color scales.

Northeastern Brazil is characterized by drought and a semiarid climate. The distribution of cases by diagnosis period varied over time, the number of cases showing progressive incidence increase: 16 cases in the 1990s (5.5%), 95 cases (32.5%) in the 2000 to 2010 period, and 180 cases (61.6%) cases between 2011 and 2021 (Fig. 3A). Since 2010, the fewest cases occurred in 2020 (5 cases), probably associated with the COVID-19 pandemic. Distribution of cases across time was uneven. In 2004 and between 2015 and 2017 there was a spike of cases, particularly in the state of Piauí (Fig. 3A). For this state, precipitation (Fig. 3B, p=0.015) and precipitation one-year prior (Fig. 3C, p=0.001) were significant predictors.

**Figure 3.**
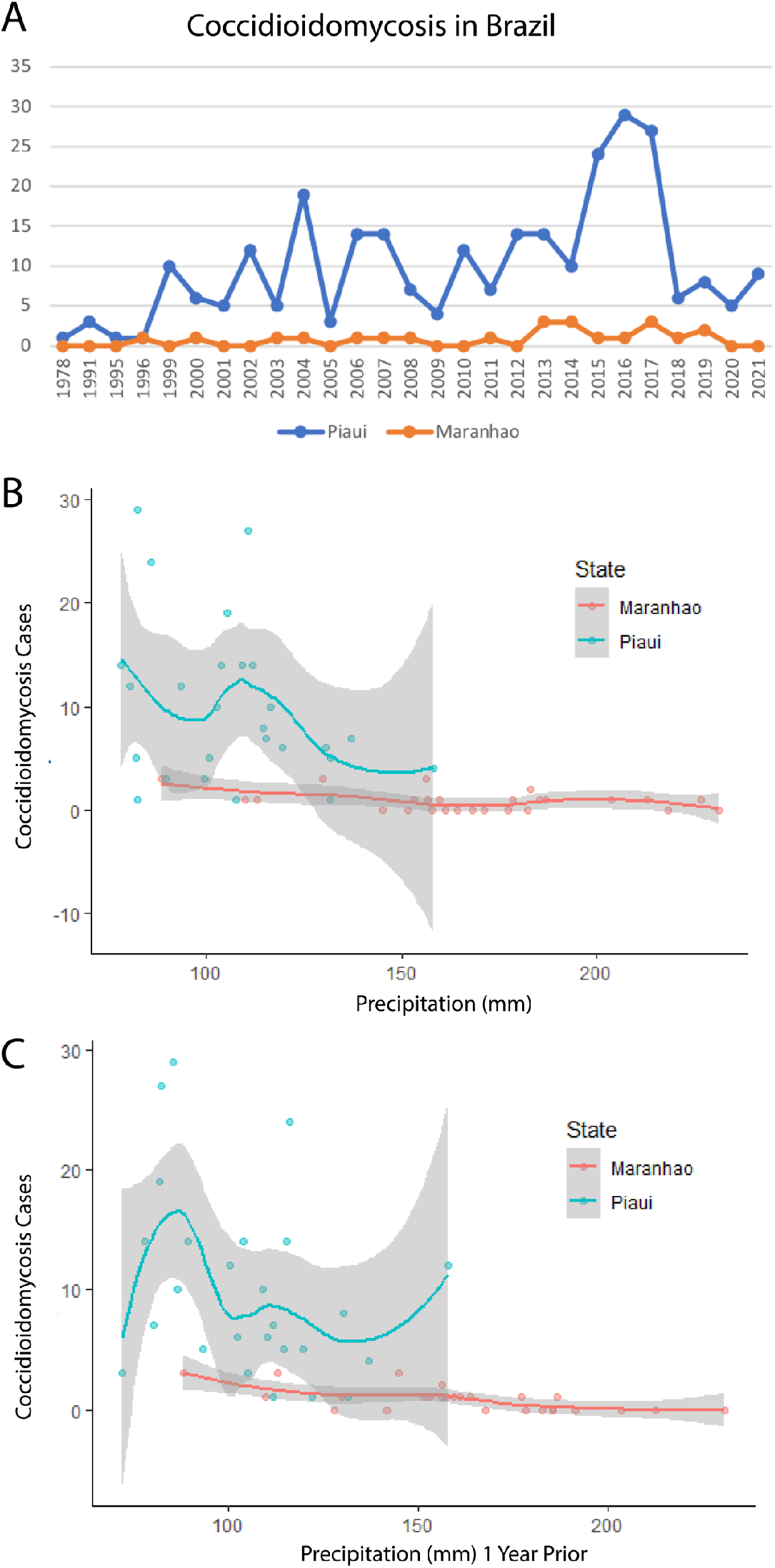
Descriptive epidemiology and climatic predictors of coccidioidomycosis in northeastern Brazil. A) Histogram shows the distribution of coccidioidomycosis cases in Piauí and Maranhão states over years. The year intervals are displayed on the x axis and case counts are displayed on the y axis. Regression analysis of precipitation (B) and precipitation one year prior (C) showing a correlation with the increase of coccidioidomycosis cases in northeastern Brazil. The volume of precipitation in displayed on the x axis in millimeters (mm) and case counts are displayed on the y axis.

There appears to be a negative relationship between high precipitation and coccidioidomycosis. When the yearly precipitation increases over 100mm, cases decrease (Fig. 3C). Temperature was not a significant predictor (p=0.87), although there does appear to be a slight positive correlation between higher temperatures and cases. The municipalities’ altitude ranged from 72m to 600m above the sea, mean temperature ranged from 24.2°C to 27.4°C, and annual mean pluviometry ranging from 527.9 mm to 1,490 mm. Analysis of physicochemical properties of soil where microepidemics occurred revealed acidic soils, with acidity ranging from medium (five sites/55.6%) to high (four sites/44.4%) (Table 3). The soil texture was sand (44.4%), clay-sand (44.4%) and clay (11.1%). Electrical conductivity, or soil salinity, was very low in the seven soils examined, being classified as “non-saline” soils; electrical conductivity in samples from the municipalities of Caridade and João Costa were not evaluated for technical reasons. The presence of organic matter was low in five soils (55.6%), medium in three (33.3%) and high in only one (11.1%). Lastly, species niche modeling suggests that the endemic area of coccidioidomycosis in Brazil is larger than previously observed. According to the logistic regression model output, suitable habitat for *Coccidioides* exists in eastern Maranhão, Piauí, and Ceará states, which represents the known endemic areas of the disease in Brazil and overlaps with our case series herein reported. However, potential coccidioidomycosis hotspots are predicted in Northern Bahia, Sergipe, Alagoas, Pernambuco, Paraiba and Rio Grande do Norte states (Fig. 4).

**Table 3:** Physicochemical properties of soil collected in areas of coccidioidomycosis outbreaks in Northeast Brazil. The location, site, pH, percentage of organic matter, texture and salinity are displayed.

| Municipality | Site | pH (acidity) | Organic matter (%) | Texture | Salinity (CE) dS/cm |
| --- | --- | --- | --- | --- | --- |
| Caridade | Armadillo burrow | 5,7<br>(medium) | 1,11<br>(low) | Clayey-sandy | Non determined |
| João Costa | Armadillo burrow | 5,2<br>(medium) | 2,17<br>(medium) | Clayey-sandy | Non determined |
| Sebastião Leal | Armadillo burrow | 4,5<br>(high) | 0,72<br>(low) | Sandy | 0,2 (non-saline) |
| Prata do Piauí | Armadillo burrow | 5,7<br>(medium) | 1,7<br>(medium) | Sandy | 0,3 (non-saline) |
| Elesbão Veloso | Armadillo burrow | 5,2<br>(medium) | 0,68<br>(low) | Clayey-sandy | 0,2 (non-saline) |
| Canto do Buriti | Armadillo burrow | 4,0<br>(high) | 6,98<br>(high) | Arenosa | 2,5 (non-saline) |
| Monsenhor Gil | Armadillo burrow | 5,2<br>(medium) | 0,95<br>(low) | Argilosa | 0,3 (non-saline) |
| Altos | Coal plant | 4,8<br>(high) | 1,66<br>(medium) | Sandy | 0,2 (non-saline) |
| Teresina | Quarry | 4,9<br>(high) | 1,02<br>(low) | Sandy | 0,1 (non-saline) |

**Figure 4.**
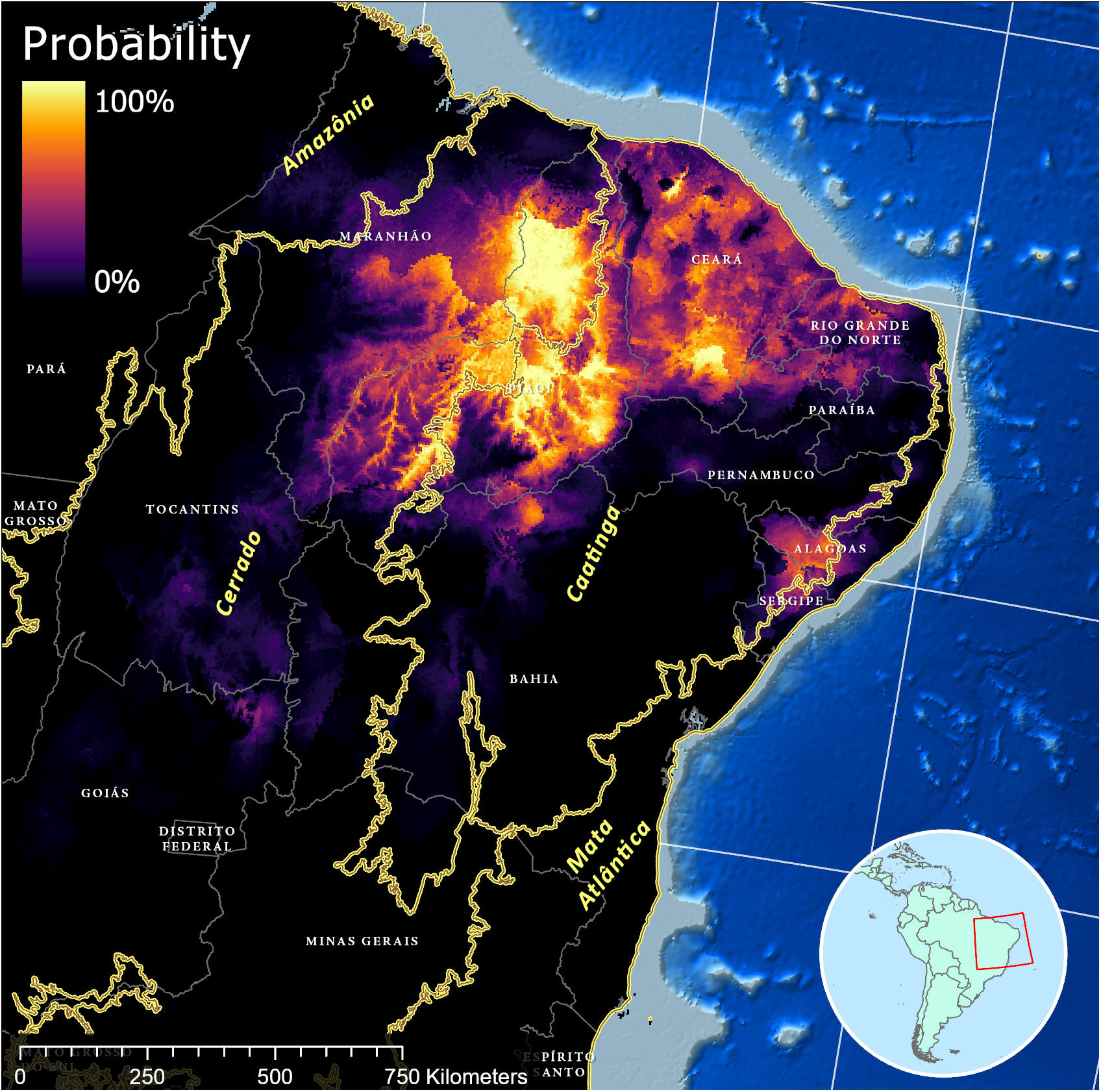
Species distribution modeling of *Coccidioides* sp. in northeastern Brazil. Species distribution modeling was conducted using environmental and ecological data to estimate the potential geographic range of Coccidioides spp. occurrence. The distribution map is based on a combination of climatic factors, data from field surveys, laboratory analysis, and previous records of *Coccidioides* spp. occurrences and considered to the modeling process. The color gradient on the map represents the probability of *Coccidioides* sp. presence, with lighter shades indicating higher likelihood. The northeastern states and the main biomes in Brazil are also delimited.

### 4 Phylogenomic and population genetics reveals a novel *C. posadasii* genotype in northeastern Brazil

Clinical isolates retrieved from human coccidioidomycosis cases in Brazil were subjected to whole genome sequencing and evolutionary analysis. The phylogenomic tree obtained recapitulates the genus *Coccidioides* harboring two district species as expected: *C. immitis* and *C. posadasii*. The 13 isolates from Brazil belong to the *C. posadasii* species, more specifically nested within the Texas/Mexico/SouthAmerica (TX/MX/SA) clade along with the previously published genome from Brazil, B5773^2^ (Fig. 5A). Within this clade, all Brazilian strains cluster in a unique branch. Importantly, strains from South America reveal divergent evolutionary trajectories. Strains from Venezuela (CARIBE clade) and Brazil (TX/MX/SA clade) arose from distinct lineages, rather than clustering together as a South American clade. Low intrinsic genetic variation is inferred from shorter branch length of Brazilian isolates compared to other taxa from the TXMXSA or ARIZONA populations indicating lower number of substitutions per site. The same pattern is observed for the VENEZUELA cluster suggesting multiple and recent introduction in South America. Moreover, the overall nucleotide diversity for Brazilian isolates is low (πχ=0.1%) compared to the TXMXSA clade (πχ=4.5%), ARIZONA (πχ=5.7%) or to GUATEMALA (πχ=1.5%) but is similar to that of VENEZUELA (πχ=0.09%).

**Figure 5.**
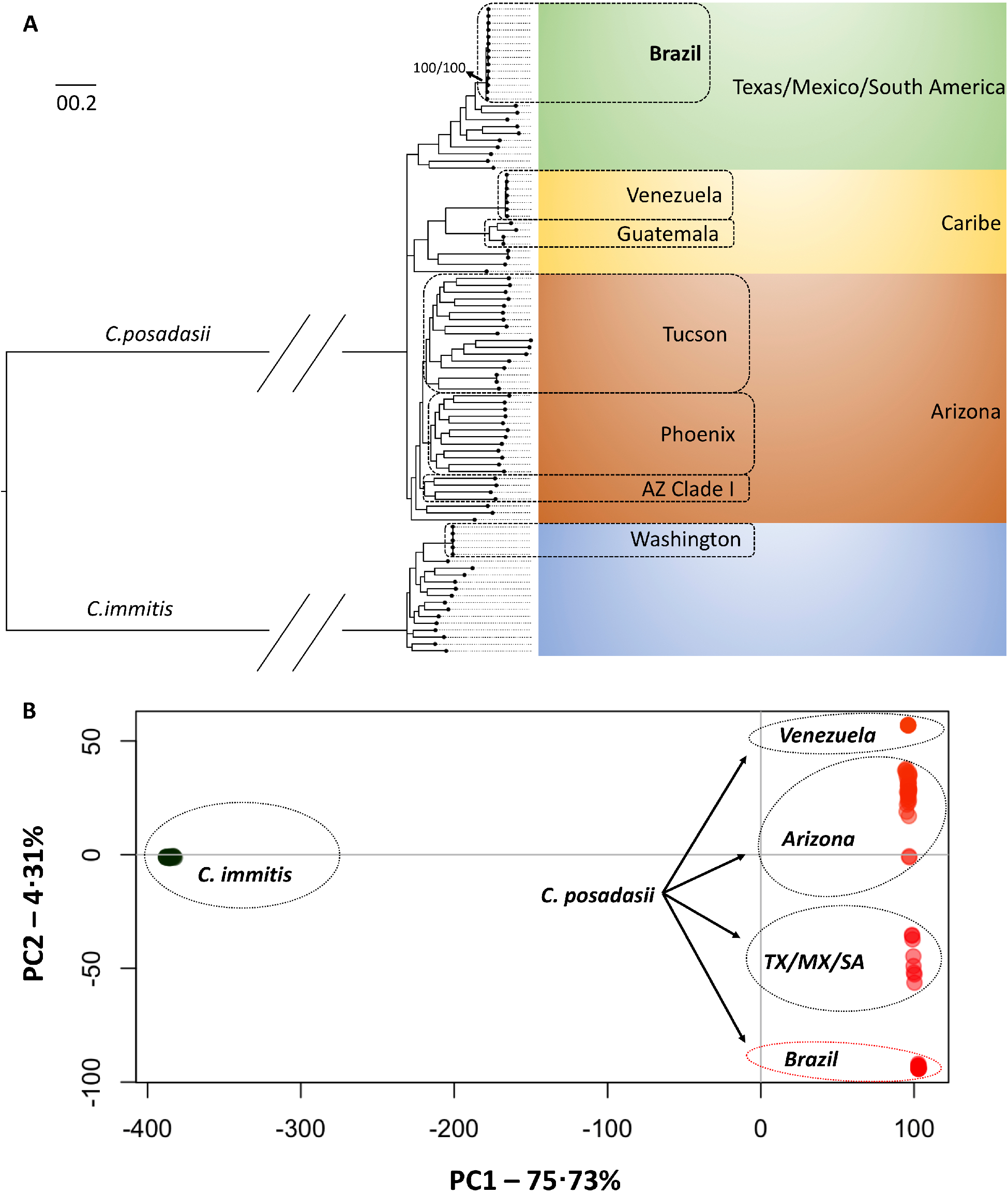
Evolutionary analysis of *Coccidioides posadasii* isolated in northeastern Brazil. A) The Maximum Likelihood phylogenomic tree shows that clinical isolates from Brazil are nested within *C. posadasii* species in a unique cluster with the Texas/Mexico/South America population. The branches are proportional to the number of mutations and 1000 ultrafast bootstraps and SH-aLRT were used to test branch support and added to the Brazil clade. The nodes in the tree represent common ancestors, and the branching points indicate the divergence of lineages. The tree was rooted with *C. immitis*. B) Principal Coordinate Analysis (PCA) shows the genetic distribution of *Coccidioides*, a fungal pathogen. These coordinates are derived from a matrix of whole-genome polymorphisms or dissimilarities between the isolates from diverse geographical locations The PCA plot depicts the distribution of the isolates along the first two principal coordinates; PC1 axis indicates the degree of genetic dissimilarity or similarity between *C. immitis* and *C. posadasii* while PC2 captures a smaller proportion of the total variation of *C. posadasii* clusters.

Next, we investigated the population distribution of the Brazilian strains compared to other *C. posadasii* individuals using PCA analysis. The PC1 corresponded to most (75.73%) of the total variation and separates *C. posadasii* from *C. immitis*. PC2, corresponded to 4.31% of the total variation and revealed distinct populations within *C. posadasii*. The Brazilian cluster is separated from the ARIZONA, CARIBE and TXMXSA groups futher supporting a unique genetic background compared to other *C. posadasii* (Fig. 5B)

## Discussion

Coccidioidomycosis was first officially reported in Brazil in 1970s from Bahia and Piauí^10,11^. However, it was not included in the geographical distribution map until 1998 when acute disease and outbreaks in Piauí and Ceará states were documented^12,14^. Presently, coccidioidomycosis is emerging in Brazil, particularly in Piauí state, where the incidence is increasing^12,15^. This study found that coccidioidomycosis primarily affects young, immunocompetent afro-descendent males who acquire the disease by inhaling dust during armadillo hunting, a common practice in the Northeastern states of Brazil (Fig. 1). Generally, persons most at risk are from highly marginalized populations and engaged in risky activities. Inhaling a high burden of arthroconidia increases the incidence of severe symptoms^16^. *Coccidioides* spp. are found in animal burrows, where they degrade organic matter derived from mammals (e.g., keratin, bones, arthropods, excreta), and are well adapted to harsh environments with high mean annual temperature and low water viability^6^.

This report is the largest case series study of the disease in South America but a key question remains unanswered: Why is the disease burden in Latin America lower than in the USA? Coccidioidomycosis is underreported due multiple factors including the lack of 1) availability of rapid, accurate, and reliable diagnostics; 2) reporting to public health authorities; 3) awareness of the disease prevalence - which might be wider than previously known^1,12,17^ (Fig. 4). Species niche modeling analysis suggests the disease might occur in other northeastern states of Brazil, where the semiarid environment could support the growth and sporulation of *C. posadasii*, and global warming models predict *C. posadasii* will expand^8^. Most cases were diagnosed after significant dust-disturbing activities that produce a high load of dust and arthroconidia, leading to acute pneumonia shortly after exposure. Thus, mild or non-pulmonary forms of the disease might be missed in the current study.

Even in areas where coccidioidomycosis is more common, frequent misdiagnosis occurs with symptoms erroneously attributed to bacterial and viral pneumonia, tuberculosis, or occupational disease^18^. Thus, there is a significant need to train clinical professionals to “think fungus” and improve diagnostics for coccidioidomycosis at a lower cost. The number of cases reported for other South American countries such as Argentina and Venezuela have long been stably low. Indeed, the low numbers cannot reflect the actual burden due to the lack of compulsory notification and diagnostic tools. Interestingly, cases in Argentina and Venezuela are predominantly classified as chronic forms, either pulmonary or disseminated, and not associated with micro-epidemics^19,20^. In fact, the disease in Brazil resembles North American outbreaks, namely the vast predominance of acute pulmonary disease arising up to 30 days after a high-exposure episode. Signs and symptoms related to acute pulmonary involvement like fever, dry cough, thoracic pain, dyspnea, asthenia, among others, overlap with those described in the USA. Also, among our 100 cases in whom more detailed data were available, only a subset of patients developed severe disease: 12% with severe pneumonia and acute respiratory failure and 7% with disseminated disease, figures that overlap with those observed for the general population in the southwestern USA, but much lower than those for some specific ethnicities such as African Americans or Filipinos^16,21^.

An analysis of predictors of severe disease or mortality such as the role of ethnicity could not be carried out due to the high rate of miscegenation in the population and because skin color phenotype does not accurately correlate with patient genetic background. The resulting mortality rate – 8%– was also higher than that reported in other large USA cohorts, e.g., those of Kern County (3%) and Naval Medical Center San Diego (0.9%)^22,23^. The reasons remain to be fully determined and are certainly multifactorial, but include diagnostic delays and access to care. Of note, the only two indigenous patients of the cohort had a complicated disease and died; however, both also had issues related to medical assistance. Patients in our cohort were treated in accordance with current guidelines, with deoxycholate amphotericin B as the choice for severe, life-threatening disease, and triazoles for the remaining patients. Only a few cases resolved spontaneously. Among azoles, itraconazole was most used, and not fluconazole, perhaps due to Brazilian clinicians’ experience in the treatment of other endemic mycoses, e.g., paracoccidioidomycosis and sporotrichosis. The overall result in our cohort is that treatment was successful, with rare failures related to patients with advanced pulmonary or disseminated disease, similar to other cohorts^24^. Dissemination to CNS was rare.

However, thoracic imaging revealed the presence of multiple nodular lesions, a finding rather uncommon in the USA cohorts^16^. This may reflect exposure to high inoculum. In fact, armadillo burrows represent a rich ecological niche supporting the growth and sporulation of *Coccidioides* spp., which degrade organic matter derived from mammals (e.g., keratin, bones, arthropods, excreta, etc.) and are well adapted to live in harsh environments with high temperatures and low water viability. Armadillo hunting necessarily involves digging deep in the burrows, and results in high inoculum exposure. This exposure may relate to the other unexpected finding of persistent alterations on thoracic imaging in almost all (92.1%) patients from a small subset that underwent a post-treatment reassessment. Despite being asymptomatic and having an otherwise normal physical examination, multiple nodules were observed, either calcified or not, which differs from other studies in which residual lesions were lower (2%-39.61%)^25,26^, and typically single nodules^27^.

Patient or environmental factors specific to Piauí and Maranhão do not explain higher frequency of diagnoses in those states, compared to other states in the Northeast. For example, the main biome of the Northeastern macroregion is the Caatinga where many of the cases were diagnosed. However, the southern parts of Piauí and Maranhão states are mostly associated with Cerrado suggesting that *C. posadasii* might also be endemic to this biome in Northeastern Brazil. We note that both biomes share physiochemical characteristics such as high temperatures and low water viability. Low annual precipitation (Precipitation and Precipitation one-year prior bioclimatic variables) was associated with spikes in cases in Brazil^28^. In addition to the absence of dust storms and earthquakes in Brazil, which are occasionally associated with increased coccidioidomycosis in North America, we have noted important unique physicochemical features of Northeastern states of Brazil. The soils sampled in areas of outbreaks are acidic and non-saline, which is the opposite to observations in endemic areas of Arizona and California which are alkaline and saline^29^. Another relevant differential characteristic of the disease between the North and South America is the pathogen genetic background. The disease in Arizona is caused by *C. posadasii* ARIZONA population while in Central America and Venezuela is caused by *C. posadasii* CARIBE population^2^. The isolates from Brazil form a unique population within *C. posadasii* TXMXSA cluster (Fig. 5). Such genetic differences might reflect specific virulence and pathogenesis traits in North America (caused by *C. posadasii* ARIZONA) relative to South America (caused by *C. posadasii* CARIBE and TXMXSA). Although coccidioidomycosis is not a notifiable disease in Texas or Mexico, the incidence of the disease appears lower than Arizona and California in spite of similar skin test positivity^30^. It is worth noting that the *C. posadasii* TXMXSA population share a common ancestor, which might contribute to a set of traits that affect ability to cause disease after infection. Whether or not different *C. posadasii* and *C. immitis* populations display altered clinical traits is unknown.

Taken altogether, we conclude that coccidioidomycosis in Northeastern Brazil has a specific infection profile associated with armadillo hunting. In Brazil, coccidioidomycosis is a neglected disease in marginalized populations, who, for cultural and/or economic reasons, engage in activities that expose them to infection, which differs from North America where a more diverse population gets infected. We also observed that specific bioclimatic variables such as low pluviosity and extensive droughts seem key to increasing the number of cases in Brazil, a finding that resonates in the context of global warming. We also show that a unique *C. posadasii* genotype is circulating in Brazil and we propose that it might display different virulence and pathogenesis traits than other *Coccidioides* genotypes. We suggest that global and federal public and occupational health authorities should support and collaborate with established local centers in these endemic regions to better understand coccidioidomycosis in South America.

## Appendix

## Inclusion criteria

Patients showing clinical respiratory signs were included, irrespective of extra-pulmonary or hypersensitivity manifestations. Ten patients were subject of previous publications ^1-6^. Inclusion criteria included: 1) Identification of mature spherules by direct microscopic examination of any patient’s sample in preparations with 10% KOH; 2) Culture of the clinical samples in Sabouraud-agar with chloramphenicol supplemented or not with cycloheximide up to 6 weeks at 30°C; positive cultures underwent microscopic identification for hyaline mycelial producing arthroconidial cells; 3) Histopathological examination of biopsies of lung or integumentary tissue with hematoxylin eosin, PAS and Grocott’s silver impregnation. The finding of mature spherules was considered a definitive diagnosis.

### Coccidioidomycosis in dogs

Dogs exposed to dust from armadillo habitats during hunting were investigated and managed according to the veterinary practices as described elsewhere^7^. Symptoms of coccidioidomycosis in dogs such as lameness, weakness, fever, fatigue, diminished appetite, weight loss, coughing, and joint discomfort, as well as pain in the back and neck were observed. Disseminated disease was also observed throughout the body since the dogs may exhibit seizures or suffer from vision loss due to the involvement of the central nervous system. The definitive diagnosis relied on isolation of the pathogen in microbiological cultures, by cytological analysis of tissue samples or direct microscopic examination of dog sputum samples in preparations with 10% KOH.

### Coccidioides niche modeling and relationship between climate and coccidioidomycosis cases in Brazil

We utilized 19 bioclimatic variables obtained from WorldClim database at a resolution of 2·5 km (www.worldclim.org/current)^8^ as predictors, testing for autocorrelation and selecting variables with r<0·80^9^. The final set of variables included Temperature Seasonality, Precipitation Seasonality, Precipitation during the coldest quarter, and Precipitation during the warmest quarter. The average final map had a logistic output, with suitability values ranging from 0 (unsuitable habitat) to 1 (suitable habitat). We employed the machine learning Maxent model (“dismo” R package ^10^ with 63 presence points and 10,063 total points to determine the Maxent distribution, resulting in an AUC of 0·87 using 1,000 maximum iterations. The model was able to discriminate between positive and negative sites for the presence of Coccidioides relatively well (AUC=0·87) and had a greater sensitivity than specificity (AUC=0·926). The average final map had a logistic output, with suitability values ranging from 0 (unsuitable habitat) to 1 (suitable habitat). To investigate the relationship between climate and coccidioidomycosis cases in Brazil ^11^, we used negative-binomial regression models (MASS R package^12^) with predictor variables of total yearly precipitation, average yearly temperature, and total precipitation of the previous year, selecting the best model with the lowest AIC. We checked for collinearity among the predictor variables in the model using a correlation matrix with the GGally R package^13^. All variables that were kept in the model had Person’s correlation coefficients <0·6.

### DNA sequencing and evolutionary analysis

Fungal growth was achieved by plating clinical specimens (see above) into the fungibiotic media Mycosel. DNA was extracted from 500mg of cells ^14^, assessed for integrity via agarose gel electrophoresis, 1μg of input DNA subjected to libraries preparation using the NEBNext® Ultra™ II DNA Library Prep Kit (New England Biolabs) and quantified via qPCR, Bionalyzer (Agilent) and Qubit (Invitrogen). DNA libraries were sequenced using the NovaSeq 6000 instrument (kit v1·5 - 300 cycles - 2×150bp) in a high-throughput mode. Initial quality control for the sequenced reads were performed using the FastQC pipeline^15^. We mapped the Illumina fastq reads into the reference *C. posadasii* strain Silveira ^16^ using bwa-mem v 0·7·7 ^17^. Next, we identified mismatch intervals and indels with help of GATK v3·3 tools RealignerTargetCreator and IndelRealigner^18^. We utilized the GATK UnifiedGenotyper, a part of GATK toolkit, using the parameter het = 0·01 to account for a haploid organism to retrieve SNPs. We used the following filters to obtain high-confident SNP calls: QD = 2 || FS_filter = 60 || MQ_filter = 30 || MQ_Rank_Sum_filter = -12·5 || Read_Pos_Rank_Sum_filter = -8 (see ^19^). We purged SNPs with less than 10X coverage, with less than 90% variant allele calls, or that were identified by Nucmer ^20^ as located in duplicated regions in the reference genome. We measured nucleotide diversity of the strains from Brazil using the Maximum likelihood composition method available in the MEGA X software ^21^. The retrieved 502,553 SNPs across 94 taxa were submitted to phylogenomic analysis using IQTREE2^22^ software using the concatenation approach. We used ModelFinder ^23^ to calculate the best nucleotide substitution model under the Bayesian Information Criterion. The best tree topology was calculated under the Maximum Likelihood criteria and branch support was assessed using both SH-like alternate Likelihood Ratio Test (aLRT^24^) and ultrafast bootstraps with 1,000 replicates^25^. The tree topology was visualized using the FigTree v1·4·4 (http://tree.bio.ed.ac.uk/software/figtree/). Principal Coordinate Analysis (PCA) was applied via the R package adegenet^26^ to evaluate population structure. The resulting Eigenvalues were used to calculate the two main prominent variations (PC1 and PC2) and to verify the population splits within *C. posadasii*.

## Data availability

Bioproject: PRJNA1000610

Biosample: SAMN36772057-SAMN36772069

SRA deposit: SRR25495556-SRR25495568

## References

1 Laniado-Laborin, R., Arathoon, E. G., Canteros, C., Muniz-Salazar, R. & Rendon, A. Coccidioidomycosis in Latin America. Med Mycol 57, S46–S55, doi:10.1093/mmy/myy037 (2019).

2 Teixeira, M. M. et al. Population Structure and Genetic Diversity among Isolates of Coccidioides posadasii in Venezuela and Surrounding Regions. mBio 10, doi:10.1128/mBio.01976-19 (2019).

3 Kandemir, H. et al. Phylogenetic and ecological reevaluation of the order Onygenales. Fungal Diversity, doi:10.1007/s13225-022-00506-z (2022).

4 Lewis, E. R., Bowers, J. R. & Barker, B. M. Dust devil: the life and times of the fungus that causes valley Fever. PLoS Pathog 11, e1004762, doi:10.1371/journal.ppat.1004762 (2015).

5 Taylor, J. W. & Barker, B. M. The endozoan, small-mammal reservoir hypothesis and the life cycle of Coccidioides species. Med Mycol 57, S16–S20, doi:10.1093/mmy/myy039 (2019).

6 Kollath, D. R., Teixeira, M. M., Funke, A., Miller, K. J. & Barker, B. M. Investigating the Role of Animal Burrows on the Ecology and Distribution of Coccidioides spp. in Arizona Soils. Mycopathologia 185, 145–159, doi:10.1007/s11046-019-00391-2 (2020).

7 Williams, S. L. & Chiller, T. Update on the Epidemiology, Diagnosis, and Treatment of Coccidioidomycosis. J Fungi (Basel) 8, doi:10.3390/jof8070666 (2022).

8 Gorris, M. E., Treseder, K. K., Zender, C. S. & Randerson, J. T. Expansion of Coccidioidomycosis Endemic Regions in the United States in Response to Climate Change. Geohealth 3, 308–327, doi:10.1029/2019GH000209 (2019).

9 Stockamp, N. W. & Thompson, G. R., 3rd. Coccidioidomycosis. Infect Dis Clin North Am 30, 229–246, doi:10.1016/j.idc.2015.10.008 (2016).

10 Gomes, O. M. et al. [Pulmonary coccidioidomycosis. 1st national case]. AMB Rev Assoc Med Bras 24, 167–168 (1978).

11 Vianna, H., Passos, H. V. & Sant’ana, A. V. [Coccidioidomycosis. Report of the 1st case in a native of Brazil]. Rev Inst Med Trop Sao Paulo 21, 51–55 (1979).

12 Cordeiro, R. et al. Coccidioidomycosis in Brazil: Historical Challenges of a Neglected Disease. J Fungi (Basel) 7, doi:10.3390/jof7020085 (2021).

13 Eulalio, K. D. et al. Coccidioides immitis isolated from armadillos (Dasypus novemcinctus) in the state of Piaui, northeast Brazil. Mycopathologia 149, 57–61 (2001).

14 Wanke, B. et al. Investigation of an outbreak of endemic coccidioidomycosis in Brazil’s northeastern state of Piaui with a review of the occurrence and distribution of Coccidioides immitis in three other Brazilian states. Mycopathologia 148, 57–67 (1999).

15 Deus Filho, A. d. Capítulo 2: coccidioidomicose. Jornal Brasileiro de Pneumologia 35, 920–930 (2009).

16 Cox, R. A. & Magee, D. M. Coccidioidomycosis: host response and vaccine development. Clin Microbiol Rev 17, 804–839, table of contents, doi:10.1128/CMR.17.4.804-839.2004 (2004).

17 Laniado-Laborin, R. Expanding understanding of epidemiology of coccidioidomycosis in the Western hemisphere. Ann N Y Acad Sci 1111, 19–34, doi:10.1196/annals.1406.004 (2007).

18 Ampel, N. M. The diagnosis of coccidioidomycosis. F1000 Med Rep 2, doi:10.3410/M2-2 (2010).

19 Campins, H. Coccidioidomycosis in South America. A review of its epidemiology and geographic distribution. Mycopathologia et mycologia applicata 41, 25–34 (1970).

20 Canteros, C. E. et al. [Coccidioidomycosis in Argentina, 1892-2009]. Rev Argent Microbiol 42, 261–268, doi:10.1590/S0325-75412010000400004 (2010).

21 Galgiani, J. N. How does genetics influence Valley Fever? Research underway now to answer this question. AZ Medicine Fall, 30–33 (2014).

22 Einstein, H. E., Catanzaro, A. & Diseases, N. F. f. I. Coccidioidomycosis: Proceedings of the 5th International Conference on Coccidioidomycosis, Stanford University, 24-27 August, 1994 : Centennial Conference. (National Foundation for Infections Diseases, 1996).

23 Crum, N. F., Lederman, E. R., Stafford, C. M., Parrish, J. S. & Wallace, M. R. Coccidioidomycosis: a descriptive survey of a reemerging disease. Clinical characteristics and current controversies. Medicine (Baltimore) 83, 149–175, doi:10.1097/01.md.0000126762.91040.fd (2004).

24 Ampel, N. M. The Treatment of Coccidioidomycosis. Rev Inst Med Trop Sao Paulo 57 Suppl 19, 51–56, doi:10.1590/S0036-46652015000700010 (2015).

25 Knoper, S. R. & Galgiani, J. N. Coccidioidomycosis. Infectious Disease Clinics of North America 2, 861–875, doi:https://doi.org/10.1016/S0891-5520(20)30232-4 (1988).

26 Bays, D. J. et al. Natural History of Disseminated Coccidioidomycosis: Examination of the Veterans Affairs-Armed Forces Database. Clin Infect Dis 73, e3814–e3819, doi:10.1093/cid/ciaa1154 (2021).

27 Drutz, D. J. & Catanzaro, A. Coccidioidomycosis. Part II. Am Rev Respir Dis 117, 727–771, doi:10.1164/arrd.1978.117.4.727 (1978).

28 Comrie, A. C. & Glueck, M. F. Assessment of climate-coccidioidomycosis model: model sensitivity for assessing climatologic effects on the risk of acquiring coccidioidomycosis. Ann N Y Acad Sci 1111, 83–95, doi:10.1196/annals.1406.024 (2007).

29 Barker, B., Tabor, J. A., Shubitz, L. F., Perrill, R. & Orbach, M. J. Detection and phylogenetic analysis of Coccidioides posadasii in Arizona soil samples. Fungal Ecol 5, 13 (2012).

30 McCotter, O. Z. et al. Update on the Epidemiology of coccidioidomycosis in the United States. Med Mycol 57, S30–S40, doi:10.1093/mmy/myy095 (2019).

## References

1 Deus Filho, A., Rocha Filho, Z. & Wanke, B. Microepidemia de coccidioidomicose em caçadores de tatu na cidade de Floriano no estado do Piauí. J. Pneumol. 26, 2 (2000).

2 Moraes, M. A., Martins, R. L., Leal, II, Rocha, I. S. & Medeiros Junior, P. [Coccidioidomycosis: a new brazilian case]. Rev Soc Bras Med Trop 31, 559–562, doi:10.1590/s0037-86821998000600009 (1998).

3 Martinez, R. Coccidioidomicose no Brasil: relato de novo caso. Rev Soc Bras Med Trop 35, 1 (2002).

4 Veras, K. N., Figueiredo, B. C. S., Martins, L. M. S., Vasconcelos, J. T. P. & Wanke, B. Coccidioidomicose: causa rara de síndrome do desconforto respiratório agudo. J Pneumol 29, 4 (2003).

5 Vianna, H., Passos, H. V. & Sant’ana, A. V. [Coccidioidomycosis. Report of the 1st case in a native of Brazil]. Rev Inst Med Trop Sao Paulo 21, 51–55 (1979).

6 Wanke, B. et al. Investigation of an outbreak of endemic coccidioidomycosis in Brazil’s northeastern state of Piaui with a review of the occurrence and distribution of Coccidioides immitis in three other Brazilian states. Mycopathologia 148, 57–67 (1999).

7 Graupmann-Kuzma, A. et al. Coccidioidomycosis in dogs and cats: a review. Journal of the American Animal Hospital Association 44, 226–235 (2008).

8 Hijmans, R. J., Cameron, S. E., Parra, J. L., Jones, P. G. & Jarvis, A. Very high resolution interpolated climate surfaces for global land areas. International Journal of Climatology 25, 1965–1978, doi:https://doi.org/10.1002/joc.1276 (2005).

9 Hernandez, P. A., Graham, C. H., Master, L. L. & Albert, D. L. The effect of sample size and species characteristics on performance of different species distribution modeling methods. Ecography 29, 773–785, doi:https://doi.org/10.1111/j.0906-7590.2006.04700.x (2006).

10 Elith, J.* et al. Novel methods improve prediction of species’ distributions from occurrence data. Ecography 29, 129–151, doi:https://doi.org/10.1111/j.2006.0906-7590.04596.x (2006).

11 Camarillo-Naranjo, J. M., Álvarez-Francoso, J. I., Limones-Rodríguez, N., Pita-López, M. F. & Aguilar-Alba, M. The global climate monitor system: from climate data-handling to knowledge dissemination. International Journal of Digital Earth 12, 394–414, doi:10.1080/17538947.2018.1429502 (2019).

12 Venables, B. & Ripley, B. (2002).

13 GGally: Extension to ‘ggplot2’ (2022).

14 Muniz Mde, M., Morais, E. S. T. P., Meyer, W., Nosanchuk, J. D. & Zancope-Oliveira, R. M. Comparison of different DNA-based methods for molecular typing of Histoplasma capsulatum. Appl Environ Microbiol 76, 4438–4447, doi:10.1128/AEM.02004-09 (2010).

15 FastQC: a quality control tool for high throughput sequence data (http://www.bioinformatics.babraham.ac.uk/projects/fastqc, 2010).

16 de Melo Teixeira, M. et al. A chromosomal-level reference genome of the widely utilized Coccidioides posadasii laboratory strain “Silveira”. G3 Genes|Genomes|Genetics 12, jkac031, doi:10.1093/g3journal/jkac031 (2022).

17 Li, H. & Durbin, R. Fast and accurate short read alignment with Burrows-Wheeler transform. Bioinformatics 25, 1754–1760, doi:10.1093/bioinformatics/btp324 (2009).

18 McKenna, A. et al. The Genome Analysis Toolkit: a MapReduce framework for analyzing next-generation DNA sequencing data. Genome Res 20, 1297–1303, doi:10.1101/gr.107524.110 (2010).

19 Teixeira, M. M. et al. Population Structure and Genetic Diversity among Isolates of Coccidioides posadasii in Venezuela and Surrounding Regions. mBio 10, doi:10.1128/mBio.01976-19 (2019).

20 Kurtz, S. et al. Versatile and open software for comparing large genomes. Genome Biol 5, R12, doi:10.1186/gb-2004-5-2-r12 (2004).

21 Stecher, G., Tamura, K. & Kumar, S. Molecular Evolutionary Genetics Analysis (MEGA) for macOS. Mol Biol Evol 37, 1237–1239, doi:10.1093/molbev/msz312 (2020).

22 Minh, B. Q. et al. IQ-TREE 2: New Models and Efficient Methods for Phylogenetic Inference in the Genomic Era. Mol Biol Evol 37, 1530–1534, doi:10.1093/molbev/msaa015 (2020).

23 Kalyaanamoorthy, S., Minh, B. Q., Wong, T. K. F., von Haeseler, A. & Jermiin, L. S. ModelFinder: fast model selection for accurate phylogenetic estimates. Nat Methods 14, 587–589, doi:10.1038/nmeth.4285 (2017).

24 Anisimova, M. & Gascuel, O. Approximate likelihood-ratio test for branches: A fast, accurate, and powerful alternative. Syst Biol 55, 539–552, doi:10.1080/10635150600755453 (2006).

25 Minh, B. Q., Nguyen, M. A. & von Haeseler, A. Ultrafast approximation for phylogenetic bootstrap. Mol Biol Evol 30, 1188–1195, doi:10.1093/molbev/mst024 (2013).

26 Jombart, T. & Ahmed, I. adegenet 1.3-1: new tools for the analysis of genome-wide SNP data. Bioinformatics 27, 3070–3071, doi:10.1093/bioinformatics/btr521 (2011).

